# EFFECT OF PROPHYLACTICALLY ADMINISTERED KETOROLAC AND DICLOFENAC POTASSIUM ON THE EFFICACY OF INFERIOR ALVEOLAR NERVE BLOCK IN PATIENTS WITH SYMPTOMATIC IRREVERSIBLE PULPITIS, DOUBLE-BLIND RANDOMIZED CONTROL TRIAL

**DOI:** 10.1101/2023.10.05.23295410

**Authors:** Sara Bano, Waleed Ishaq, Sadaf Islam, Sara Hamdani, Sana Ashfaq, Amna Moghal

## Abstract

**Background:** Inferior alveolar nerve block is the most frequently used local anesthetic agent, administered to achieve regional anesthesia for root canal treatment, however, in cases of irreversible pulpitis, there may be anesthetic resistance. To overcome this issue, many conservative and invasive supplementary procedures are introduced to increase the efficacy of nerve block, including prophylactic use of orally administered NSAIDs.

**Objective:** To compare the effect of prophylactically administered Ketorolac and Diclofenac Potassium on the efficacy of inferior alveolar nerve block in patients presenting with symptomatic irreversible pulpitis.

**Methodology:** This double-blind study included a total number of 130 patients with a diagnosis of symptomatic irreversible pulpitis which was confirmed with a cold test. Before initiating the treatment, the patients were asked to rate their pain on a VAS with pain divided into four categories, no pain, mild pain, moderate pain, and severe pain. 10 mg Ketorolac and 50 mg Diclofenac potassium were equally divided into two groups and 1 tablet of each was orally administered to patients 1 hour before initiating root canal treatment. IANB was given and a root canal procedure was initiated under full aseptic protocol. The pain score was recorded on VAS during endodontic access preparation and root canal instrumentation. Data was analyzed through cross-tabulation and the Chi-square test was applied. (*P* value = 0.05)

**Results:** The comparison of prophylactically administered ketorolac and diclofenac potassium on the efficacy of inferior alveolar nerve block depicted the success rate was 64.6% in Ketorolac group as compared to 43.1% in Diclofenac potassium group.

**Conclusion:** Oral premedication with NSAIDs can improve the efficacy of Inferior alveolar nerve block in a non-invasive manner with better results yielded with the use of orally administered Ketorolac as compared to Diclofenac Potassium.

## INTRODUCTION

The pain of dental origin is excruciating in nature and its management is the most sought-after component of clinical dentistry. One of the major causes of dental pain, irreversible pulpitis, is usually treated through root canal therapy, which itself is quite a painful procedure.^1^ This limitation of the endodontic procedure is overcome by administering intraoral local anesthesia. For achieving regional anesthesia in mandibular molars, Inferior alveolar nerve block (IANB) is the most frequently used injection technique. IANB is usually effective in healthy pulp; however, the technique fails in 30-90% of cases with an inflamed pulp.^2,3^

Multiple techniques have been introduced to overcome this anesthetic resistance of teeth with irreversible pulpitis including the use of different anesthetic drugs and the secondary administration of anesthesia by invasive approaches such as intra-osseous or ligamentary injection techniques.^4,5^ Despite approved success rates, the use of intra-osseous and ligamentary approaches has been limited due to their invasiveness and the use of cumbersome technical instruments with a risk of injuring the dental root and causing temporary tachycardia.^6^ To overcome this problem, prophylactic use of analgesics or anxiolytics has been suggested in the literature. Nonsteroidal anti-inflammatory drugs (NSAIDs) are the most commonly used analgesics for dental pain.^7^

NSAIDs inhibit cyclooxygenase (COX) enzyme which is responsible for the transformation of arachidonic acid into prostaglandins and thromboxanes, thereby exerting anti-inflammatory action. NSAIDs exhibit their analgesic effect, indirectly, through the reduction of inflammation, and, directly, due to action upon the central nervous system (CNS).^8,9^

The two most frequently used nonsteroidal anti-inflammatory drugs (NSAIDs), Diclofenac and ketorolac are used to manage moderate to severe dental pain, which can be administered orally, intravenous, intranasal, and through the use of transdermal patches.^10–13^ These drugs can non-selectively inhibit the COX enzyme pathway. Since an ample number of teeth indicated for root canal treatment are diagnosed with acute irreversible pulpitis, these NSAIDs may improve the efficacy of IANB. Due to their short half-life and ability to be immediately released, they would be considered ideal to use in a single dosage prior to the endodontic therapy in cases with symptomatic irreversible pulpitis.^14^ When administered orally, they can provide adequate clinical efficacy with convenience, as compared to the use of intramuscular or intravenous injection, which might lead to discomfort and fear and is not well accepted by some patients.^15^

The effects of premedication with a variety of drugs administered 30 minutes to 1 hour before commencement of endodontic treatment of patients with symptomatic irreversible pulpitis have been widely studied, and mixed results have been reported. A systematic review by *Shirvani et al*. recommended the use of a single dose of certain types of NSAID’s to maximize the therapeutic efficacy of the anesthetic.^16^ In another study, the success of IANB in patients with irreversible pulpitis was compared 1 hour after administration of ketorolac and ibuprofen, and no significant effect was reported.^17^ *Saha et al*. evaluated the efficacy of premedication with ketorolac, diclofenac potassium and cellulose powder used as a placebo, 1 hour prior to administration of IANB and found out that post-injection VAS Score was least in ketorolac group followed by Diclofenac group and it was maximum in the placebo group.^18^ Therefore, the objective of this study was to compare prophylactic ketorolac and diclofenac potassium on the efficacy of inferior alveolar nerve block in patients presenting with symptomatic irreversible pulpitis and provide a better alternative to dental practitioners for effective management of pain during endodontic procedures through orally administered prophylactic medication instead of invasive accessory anesthetic techniques.

## METHODOLOGY

After taking approval from the ethical review committee of our institute under IRB Number 90/Trg – ABP1K2, this study was conducted in the Department of Operative Dentistry at Armed Forces Institute of Dentistry from 02 May, 2022 to 14 Dec, 2022. Sample size was calculated with WHO calculator, using the test for two independent proportions, with the anticipated population proportion of Ketorolac group as 0.76 and Diclofenac potassium as 0.54, keeping the power of test at 80% and significance level 5%.^18^ A sum total of one hundred and thirty healthy-looking male and female patients reporting to the Operative Dentistry Department, Armed Forces Institute of Dentistry with moderate or severe pain in a first or second mandibular molar were invited for participation in the study by the principal investigator. The age range of included patients was between 18-60 years and they were divided into three groups, Group I (18-32 years), Group II (33-46 years), and Group III (47-60 years). A definitive diagnosis of SIP (Symptomatic Irreversible Pulpitis) was made after pulp vitality status was assessed with the cold test. All patients were instructed not to take any medication 12 hours prior to treatment. Patients who gave a history of active peptic ulcers, pregnancy, lactation, allergy to NSAIDs and lignocaine, or those who were not able to give informed consent or reported mild pain according to VAS were excluded from the study. Similarly, those teeth that were necrotic during the investigation with cold test were also not included in the study. After taking informed written consent, a thorough medical and dental history of each patient was recorded, and the pulpal and periapical status of the tooth to be treated was evaluated with the help of a periapical radiograph, palpation, percussion, and cold test. Patient’s age, gender, and tooth being treated were also recorded along with, in performa. Before initiating the treatment, the patients were asked to rate their pain on a Visual analogue scale (VAS) with markings on a line indicating ten levels of pain. The scale was categorized into four groups: no pain (corresponding to 0); mild pain (corresponding to 1-3); moderate (corresponding to 4-7); and severe pain (corresponding to 8-10). A trained dental hygienist divided the 130 tablets of each NSAID into two bottles: Ketorolac 10 mg (Tab. Kelac, Rotex Medica Pharmaceuticals) and Diclofenac Potassium 50 mg (Tab. Caflam, Novartis Pharma). The bottles were masked with an opaque label and randomly assigned as Group A and B respectively.

The already diagnosed and selected patients were randomly divided into two groups through block randomization by the reception staff, while keeping the principal investigator blinded. Ketorolac group (Group 1) and Diclofenac Potassium group (Group 2) contained 65 patients each and one tablet was given to each one of the patients 1 hour before the procedure. After 1 hour of oral administration of the tablets, all patients received standard IANB injections using 1.8 mL of 2% lidocaine containing 1: 200 000 epinephrine (Septodont) administered by the principal investigator. The solution was deposited using self-aspirating syringes with 27-gauge needles (Septodont) at a rate of 1 mL/ min. Each patient was asked about tingling sensation in the lip, fifteen minutes after the administration of the IANB. In cases where profound lip numbness was not recorded within 15 min, the block was considered unsuccessful and such patients were excluded from the study. The tooth in question was then tested again with cold spray and the possible outcomes were recorded. In case of a positive response with cold spray, the test was recorded as a failure and supplemental anesthesia was given. After complete isolation with a rubber dam, standardized endodontic access was attained with safe-ended tapered diamond bur with water coolant. Pain score was recorded on the Visual Analogue Scale during access cavity preparation and root canal instrumentation. In case of moderate and severe pain during the treatment, the outcome was recorded as failure and supplemental anesthesia was given while the success of IANB was defined as no pain or mild pain based on the visual analogue scale readings during endodontic access preparation and root canal instrumentation. None of the patients reported any side effects to the drugs used.

## DATA ANALYSIS

The data was recorded and analyzed on SPSS (Version 23). Descriptive statistics were calculated for both qualitative and quantitative variables. Mean and standard deviation were calculated for quantitative variables like age, Pre and Post VAS pain score. Frequency and percentages were calculated for qualitative variables like age groups, gender, and efficacy of anesthesia. Both drug groups (Ketorolac and Diclofenac potassium) were cross tabulated with the efficacy of anesthesia of IANB and a Chi-square test was applied. Effect modifiers like age groups and gender were considered by stratification. Post-stratification Chi-square test was also applied. *P* value of 0.05 or less was considered statistically significant.

## RESULTS

The study comprised of 130 patients, randomly divided into two equal drug groups i.e., Ketorolac group and Diclofenac potassium. Both drug groups contained 65 patients each. The mean age in the ketorolac group was 39.31 ± 10.92 in contrast to 37.51 ± 10.73 in the diclofenac group. The mean Pre-VAS score in both groups was quite similar. In the ketorolac group, it was found to be 6.66 ± 1.98, in the diclofenac group it was 6.69 ± 1.85. On the contrary, substantial differences were observed in the mean Post VAS score among the two groups. It was found to be 2.69 ± 2.29 in the ketorolac group as compared to 3.95 ± 2.10 in the diclofenac group depicting more efficient pain control in ketorolac group. The cumulative descriptive statistics are presented in Table 1.

**Table 1:** Descriptive Statistics.

| Variables | N = 130 |  |  |  |
| --- | --- | --- | --- | --- |
|  | Minimum | Maximum | Mean | Std. Deviation |
| Age | 19 | 59 | 38.41 | 10.821 |
| Pre VAS Score | 4 | 10 | 6.68 | 1.910 |
| Post VAS Score | 0 | 9 | 3.32 | 2.280 |

The comparison of prophylactically administered ketorolac and diclofenac potassium on the efficacy of inferior alveolar nerve block is depicted in Table 2. The higher number of successful IANB in the ketorolac group (64.6%) as compared to the diclofenac group (43.1%) shows better results were obtained with prophylactic use of ketorolac drug. The *P* value of 0.14 depicts statistically significant results on the chi-square test.

**Table 2.** Crosstabulation efficacy of anesthesia with drug type.

|  |  | Drug type |  | Total | P value |
| --- | --- | --- | --- | --- | --- |
|  |  | Ketorolac | Diclofenac Potassium |  |  |
| Efficacy of anesthesia | Yes | 42(64.6%) | 28(43.1%) | 70(53.8%) | 0.014 |
|  | No | 23(35.4%) | 37(56.9%) | 60(46.2%) |  |

\*Confidence Interval = 95%

Data was stratified according to gender and a chi-square test was applied as shown in Table 3. Post-stratification Chi-square revealed a statistically significant association in males while females exhibited insignificant association between drugs and efficacy of anesthesia.

**Table 3.** Impact of gender on efficacy of anesthesia in both drug groups.

| Efficacy of Anesthesia | Males |  | Females |  |
| --- | --- | --- | --- | --- |
|  | Keto | Diclo | Keto | Diclo |
| Yes | 26(66.7%) | 15(40.5%) | 16(61.5%) | 13(46.4%) |
| No | 13(33.3%) | 22(59.5%) | 10(38.5%) | 15(53.7%) |
| Significance | $P = 0.022$ | | $P = 0.266$ | |

\* Confidence Interval = 95%

Based upon age groups, post-stratification chi-square test revealed that the effectiveness of anesthesia was significantly associated with drug groups within Group II (33-46 years) only, while Group I (18-32 years) and Group III (47-60 years) showed an insignificant association between both variables. The data is illustrated in Table 4.

**Table 4.**
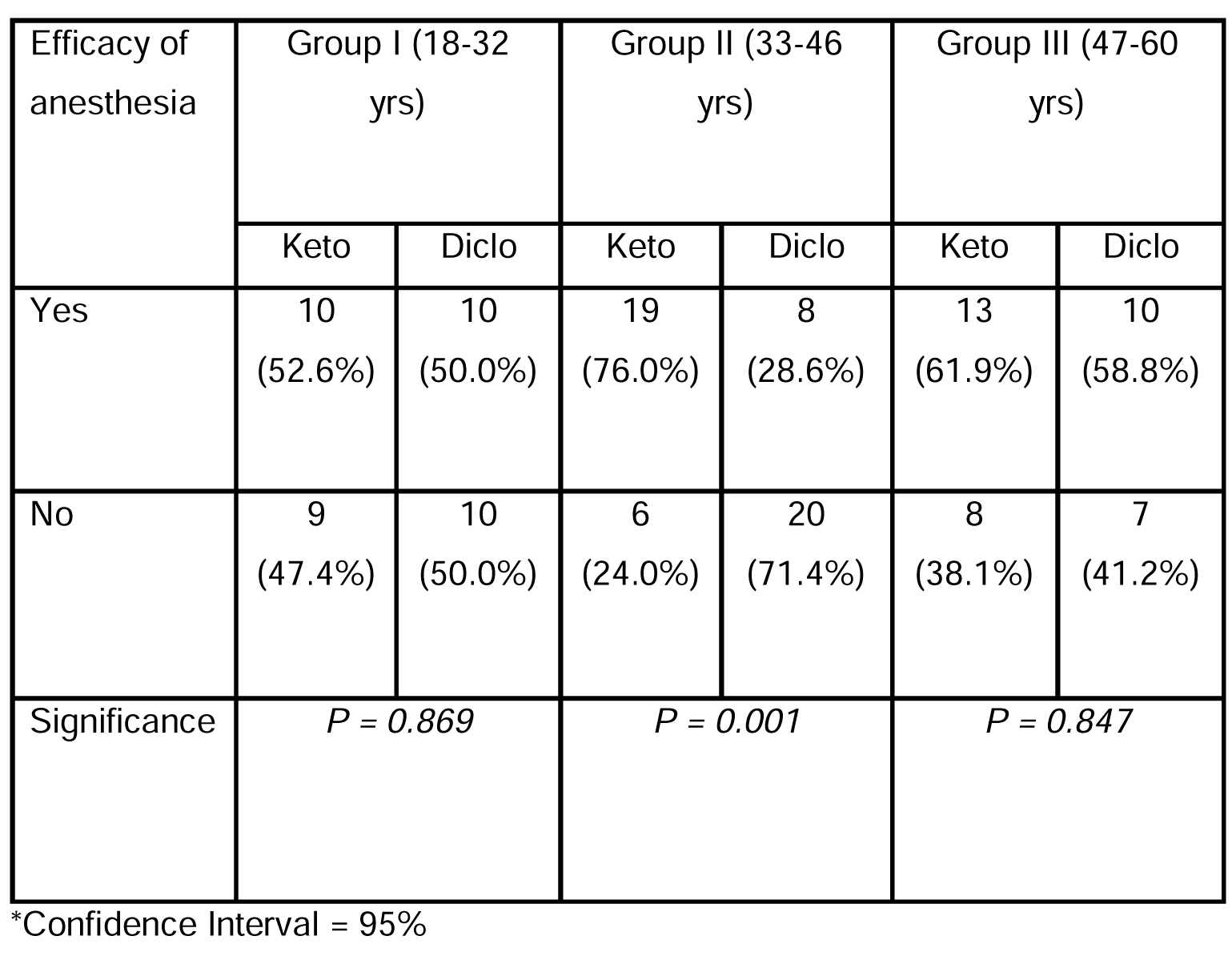
Impact of age groups on efficacy of anesthesia in both drug groups.

## DISCUSSION

Pain management is of vital importance in endodontics and local anesthesia is considered to be the primary method to achieve it. According to the literature, in the absence of pulpal inflammation, inferior alveolar nerve block is effective in approximately 96% of the cases, ^19^ but in cases of irreversible pulpitis, the success rate is greatly reduced to as low as 20-30%. ^20,21^ Anatomical variations, acute tachyphylaxis, effect of inflammation on local tissue pH, blood flow, nociception, psychological factors, and central sensitization are among the listed causes of failure to achieve anesthesia after IANB.^22^ To alleviate patient’s pain and increase compliance, alternative approaches are also under consideration by many clinicians around the world.^23^

Among the most popular techniques, oral premedication with NSAIDs stands out, as it is clinically convenient and provides promising results with mild to moderate side effects. An array of these drugs like ketorolac, ibuprofen, diclofenac potassium, diclofenac sodium, and acetaminophen group, have been studied due to their role in increasing the efficacy of IANB by blocking the COX pathway, hence, inhibiting pain sensations.^24,25^

NSAIDs act by inhibiting cyclooxygenase enzyme which is responsible for the transformation of arachidonic acid derived from linoleic acid into eicosanoids (prostaglandins and thromboxanes). Arachidonic acid gets released as a result of mechanical, chemical, or thermal insult and the resulting metabolites (prostaglandins and thromboxanes) exert effects on all body organs and tissues through potent vasodilation, resulting in increased vascular permeability, with the extravasation of fluids and white blood, resulting in inflammation. Hence, the inhibition of cyclooxygenase synthesis (COX-1 and COX-2) exerts a clear anti-inflammatory effect. COX-1 is an isoenzyme of cyclooxygenase associated with general homeostasis and is found in most organs and tissues. The analgesic effect of NSAIDs results indirectly from the reduction of inflammation, and also directly due to action upon the central nervous system (CNS).^26^

Effects of different drugs as premedication for pain relief during endodontics have been studied, however, NSAIDs are by far considered superior to other drugs, except for dexamethasone, which provides the most significant results as supported by the literature.^27^ In this study, ketorolac and diclofenac potassium were the trial drugs used, as both of these drugs provided potent pain reduction with minimal side effects.^18,28,29^ A placebo controlled study by *Kaladi et al*, concluded that ketorolac 20mg when administered 1 hour prior to treatment resulted in superior efficacy in the reduction of pain as compared to ibuprofen 400mg and placebo during and after attaining access to the pulp chamber and during canal preparation.^30^ In another study conducted by *Singh et al*, patients showed increased efficacy in IANB after premedication with ketorolac (71.4%) as compared to ibuprofen, a combination of ibuprofen and acetaminophen, and placebo.^31^ In a double-blinded controlled trial by *Praveen et al*, pain control was assessed at 6 and 12-hourly intervals after using ketorolac and prednisolone. After 6 hours, pain was relieved in the ketorolac group, however, prednisolone took more time to achieve profound results. It was attributed to better pharmacodynamics, as peak absorption levels were recorded within 2-3 hours and its effect lasted for 6 hours.^15^ Ketorolac has shown promising results in most studies, however *Kumar et al* reported statistically insignificant difference in pain improvement threshold between prophylactic ketorolac and paracetamol on the success of IANB.^32^

A systematic review by *Sivaramakrishnan et al* included four studies to evaluate the success of IANB in conjunction with ketorolac as compared to other NSAIDs and placebo. The authors concluded that oral premedication with ketorolac should be considered before initiation of endodontic treatment. ^17,28,33,34^

Diclofenac potassium, a potent NSAID, was studied by *Wali et al*, as premedication reported a 75% success rate which was inferior as compared to piroxicam which had a success rate of 90%.^35^ Another study by *Prassana et al*, was conducted to determine the effect of the administration of pre-operative lornoxicam and diclofenac potassium on the success of inferior alveolar nerve blocks in patients with irreversible pulpitis in a double-blind randomized controlled trial. In the lornoxicam group success rate was slightly high (71.4%) as compared to the diclofenac potassium group (53.5%), however, no statistically significant difference was reported between the two drug groups.^36^ *Al-Rahwani et al*, described the maximum reduction in pain at 48 hrs after premedication with 50 mg of Diclofenac potassium in a placebo controlled clinical trial.^14^ *Nagendrababu et al*, suggested in a systematic review that Ibuprofen 400 mg should be considered the standard premedication to increase the efficacy of IANB in endodontics. Alternate medications include Ketorolac and Diclofenac potassium.^29^

In a study conducted by *Saha et al*, a comparison of efficacy of IANB is made after premedication with ketorolac, diclofenac potassium, and placebo on 150 adult patients with symptomatic irreversible pulpitis in mandibular molars. The results of that study demonstrated, the success of IANB was significantly higher in patients pre-medicated with KETO when compared to DP or PLAC (*P* < 0.005), with 76.19% success IANB in patients premedicated with ketorolac, 54.76% for diclofenac group and 28.57% for placebo. ^18^ Similar findings have been replicated in the present study where we compared the efficacy of ketorolac and diclofenac potassium as pre-medications and the impact of age and gender as effect modifiers. The higher number of successful IANB in ketorolac group (64.6%) as compared to diclofenac group (43.1%) showed better results associated with prophylactic use of ketorolac drug, with better tolerance and little to no side effects, hence making it a more effective analgesic. Upon investigating the impact of effect modifiers, it was determined that male gender and middle age group (33-46 years) had significantly better efficacy of anesthesia with ketorolac.

## CONCLUSION

Oral premedication with NSAIDs like Ketorolac and Diclofenac potassium when given 1 hour prior to administration of inferior alveolar nerve block has been found to help reduce pain intensity, resulting in an effective nerve block. Oral pre-medication with ketorolac may result in a significantly higher percentage of successful inferior alveolar block in patients with irreversible pulpitis as compared to pre-medication with Diclofenac potassium.

### Limitations

The study included a relatively small sample size of 130 patients, which may limit the generalizability of the findings. A larger sample size would have provided more robust results. There has been ample data on the comparison of different NSAID’s but the lack of comparison of Ketorolac and Diclofenac potassium, made it difficult to relate to the previous studies. Another limitation of this study was the unequal distribution of first and second mandibular molars, which was included to improve the total sample size.

## Acknowledgement

The authors thank Dr. Hannan Humayun Khan at Armed Forces Institute of Dentistry and Dr. Qurat ul Khan from Shifa College of Dentistry for their utmost guidance and support in conduction of this study.

## Trial registry number

ACTRN12622000633785 (ANZCTR)

## Data Availability Statement

The datasets analyzed are available in the data repository through: Bano, Sara; Ishaq, Waleed (2023), “Effect of prophylactically administered ketorolac and diclofenac potassium on the efficacy of inferior alveolar nerve block in patients with symptomatic irreversible pulpitis, a double blind randomized controlled trial “, Mendeley Data, V1, doi: 10.17632/gkr53hmkmk.1

## Declaration of patient consent

The authors of this study have taken informed consent from all participants after explaining the procedure to the patients. All efforts will be put to maintain the anonymity of patients by the author; however, it cannot be guaranteed.

